# Characterization of SARS-CoV-2 genetic structure and infection clusters in a large German city based on integrated genomic surveillance, outbreak analysis, and contact tracing

**DOI:** 10.1101/2021.02.13.21251678

**Authors:** Andreas Walker, Torsten Houwaart, Patrick Finzer, Lutz Ehlkes, Alona Tyshaieva, Maximilian Damagnez, Daniel Strelow, Ashley Duplessis, Jessica Nicolai, Tobias Wienemann, Teresa Tamayo, Malte Kohns Vasconcelos, Lisanna Hülse, Katrin Hoffmann, Nadine Lübke, Sandra Hauka, Marcel Andree, Martin P. Däumer, Alexander Thielen, Susanne Kolbe-Busch, Klaus Göbels, Rainer Zotz, Klaus Pfeffer, Jörg Timm, Alexander T. Dilthey, German COVID-19 OMICS Initiative (DeCOI)

**Author notes:** contributed equally (Andreas Walker, Torsten Houwaart, Patrick Finzer, Lutz Ehlkes).

## Abstract

Viral genome sequencing can address key questions about SARS-CoV-2 evolution and viral transmission. Here, we present an integrated system of genomic surveillance in the German city of Düsseldorf, combining a) viral surveillance sequencing, b) genetically based identification of infection clusters in the population, c) analysis of hospital outbreaks, d) integration of public health authority contact tracing data, and e) a user-friendly dashboard application as a central data analysis platform. The generated surveillance sequencing data (n = 320 SARS-CoV-2 genomes) showed that the development of the local viral population structure from August to December 2020 was consistent with European trends, with the notable absence of SARS-CoV-2 variants 20I/501Y.V1/B.1.1.7 and B.1.351 until the end of the local sampling period. Against a background of local surveillance and other publicly available SARS-CoV-2 data, four putative SARS-CoV-2 outbreaks at Düsseldorf University Hospital between October and December 2020 (n = 44 viral genomes) were investigated and confirmed as clonal, contributing to the development of improved infection control and prevention measures. An analysis of the generated surveillance sequencing data with respect to infection clusters in the population based on a greedy clustering algorithm identified five candidate clusters, all of which were subsequently confirmed by the integration of public health authority contact tracing data and shown to be represent transmission settings of particular relevance (schools, care homes). A joint analysis of outbreak and surveillance data identified a potential transmission of an outbreak strain from the local population into the hospital and back; and an in-depth analysis of one population infection cluster combining genetic with contact tracing data enabled the identification of a previously unrecognized population transmission chain involving a martial arts gym. Based on these results and a real-time sequencing experiment in which we demonstrated the feasibility of achieving sample-to-turnaround times of <30 hours with the Oxford Nanopore technology, we discuss the potential benefits of routine ultra-fast sequencing of all detected infections for contact tracing, infection cluster detection, and, ultimately, improved management of the SARS-CoV-2 pandemic.

## Introduction

SARS-CoV-2, a pandemic coronavirus first detected in late 2019 [Wu et al. 2020, Zhou et al. 2020], has infected >108 M individuals and led to >2.3 M associated deaths [World Health Organization 2021]. Until the wide availability of vaccines, non-pharmaceutical interventions to limit SARS-CoV-2 transmission will continue to play an important role in pandemic management. Genomic epidemiology [Gardy and Loman 2018, Grubaugh et al. 2019], i.e. the application of modern genomic technologies to characterize viral transmission chains [Quick et al. 2016, Grubaugh et al. 2017, Gonzalez-Reiche et al. 2020, Gudbjartsson et al. 2020, Lu et al. 2020] can crucially contribute to the design and evaluation of viral containment strategies. Its possible applications include the targeted investigation of putative outbreaks e.g. in hospitals [MacFadden et al. 2018] and care homes, as well as untargeted “surveillance sequencing” to monitor transmission dynamics and viral evolution in the population at large. In an integrated genomic epidemiology approach, the joint analysis of surveillance, outbreak and contact tracing data can enable the improved analysis of infection chains in the population and healthcare settings [Meredith et al. 2020]. While some countries have implemented national genomic epidemiology efforts for SARS-CoV-2 [COVID-19 Genomics UK (COG-UK) consortium 2020], no such programme existed in Germany until January 2021.

In summer 2020, we therefore established an integrated SARS-CoV-2 genomic epidemiology system in Düsseldorf, the capital of North Rhine Westphalia, a city of about 600,000 inhabitants in Germany’s largest metropolitan area. Our approach included untargeted longitudinal surveillance sequencing, implemented in collaboration with a large commercial diagnostic laboratory, analysis of putative SARS-CoV-2 outbreaks from the city’s largest hospital, the integration of local public health authorities, and the development of a user-friendly dashboard for the visualization of outbreaks in the context of local sequence diversity (Figure 1). Like in our analysis of Germany’s first superspreading event in Heinsberg [Walker et al. 2020], sequencing was mostly implemented with the Oxford Nanopore technology, potentially enabling rapid turnaround times.

**Figure 1:**
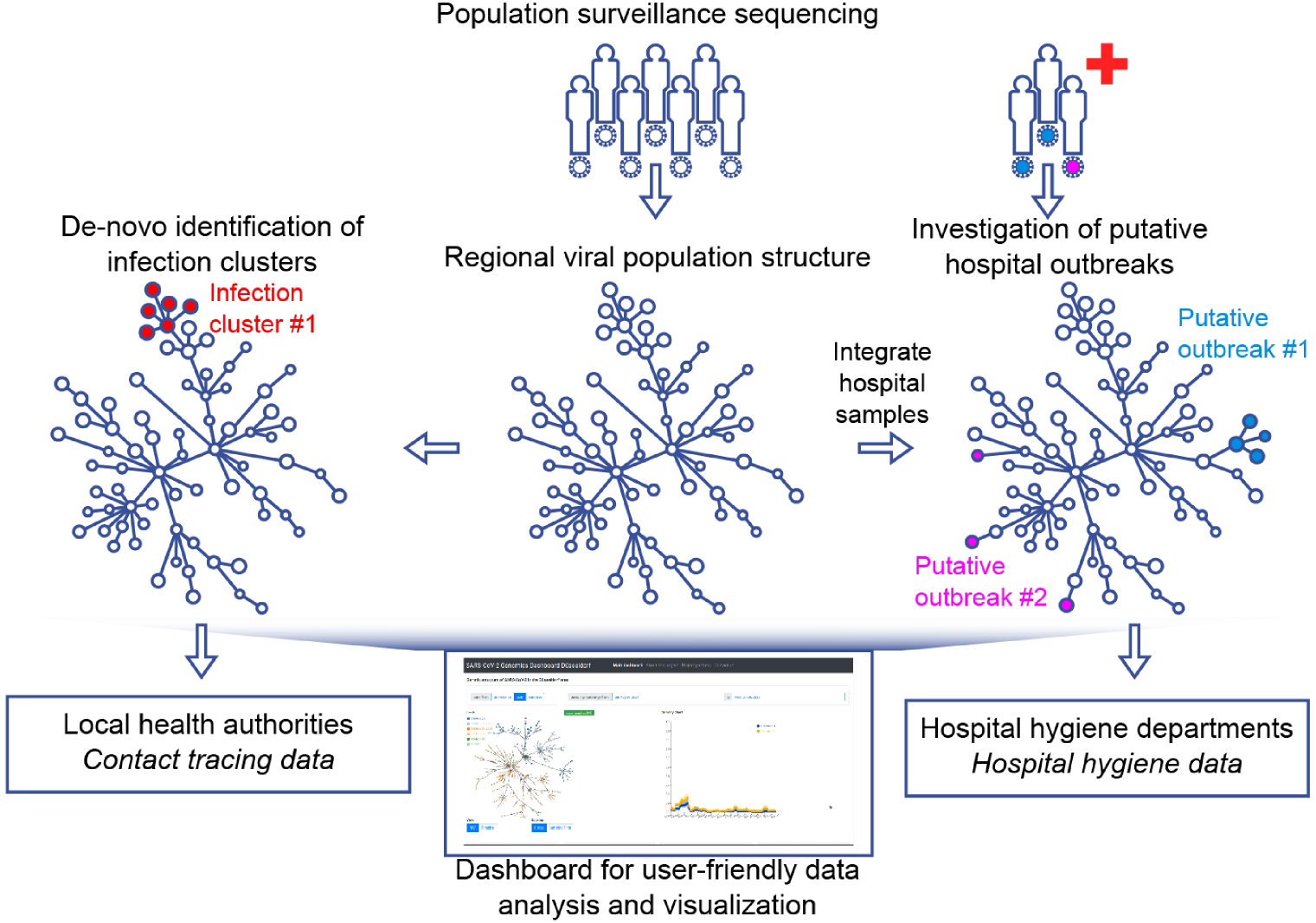
Integrated genomic surveillance in the Düsseldorf area. Population surveillance sequencing enables the characterization of local SARS-CoV-2 population structure, facilitating the discrimination between clonal hospital outbreaks (here: Putative outbreak #1) or simultaneously detected but unrelated SARS-CoV-2 hospital ward cases (here: Putative outbreak #2). Viral population surveillance data can also enable the *de novo* identification of infection clusters in the population based on the genetic data. The added value of genomic surveillance is maximized when genetic data is integrated with complementary epidemiological data or approaches, such as contact tracing or hospital outbreak data. The utilization of viral genetic data by diverse stakeholders is facilitated by providing a user-friendly real-time web application (“dashboard”) for analysis and visualization of the generated viral genomes.

## Results

### Genomic Surveillance in the Düsseldorf Region

In collaboration with a local diagnostic lab and employing a convenience sampling approach, we obtained 320 high-quality SARS-CoV-2 viral isolate genomes from samples collected in Düsseldorf between August and December 2020 (median: 19 samples / week). The collected genomes represented 0.3% of 10,276 newly diagnosed PCR-confirmed cases during the sampling period, and the proportion of sequenced cases on a weekly basis varied between 0% and 20%; during the last four weeks of the sampling period, the proportion of sequenced cases stabilized between 2% and 3%. Sequencing data, sample metadata and assembly quality are summarized in Supplementary Table 1 and Supplementary Table 2. By sample genome inclusion criteria (see Methods), all included isolate genomes were of high quality (<3000Ns). 80/320 isolate genome consensus sequences contained at least one ambiguous character (average: 0.48 ambiguous characters / genome), indicating potential intra-patient strain variability.

The development of viral population structure was largely consistent with the developments in Europe at large; for example, while clade 20E was initially found at low frequencies, it accounted for nearly half of the sequenced genomes towards the end of the sampling period (Figure 2C). One notable exception was the absence of clade 20I/501Y.V1 (equivalent to B.1.1.7 in Pangolin nomenclature; https://github.com/cov-lineages/pangolin), which had already reached significant frequencies in some European countries by the end of December 2020 (e.g., 48% in the UK or 15% in the Netherlands); the viral variant B.1.351 was also not detected. Roughly consistent with global mutation fixation rates, the number of variants (substitutions) per genome had increased to ∼20 per genome towards the end of the sampling period, representing the ongoing accumulation of viral mutations (Figure 2D).

**Figure 2:**
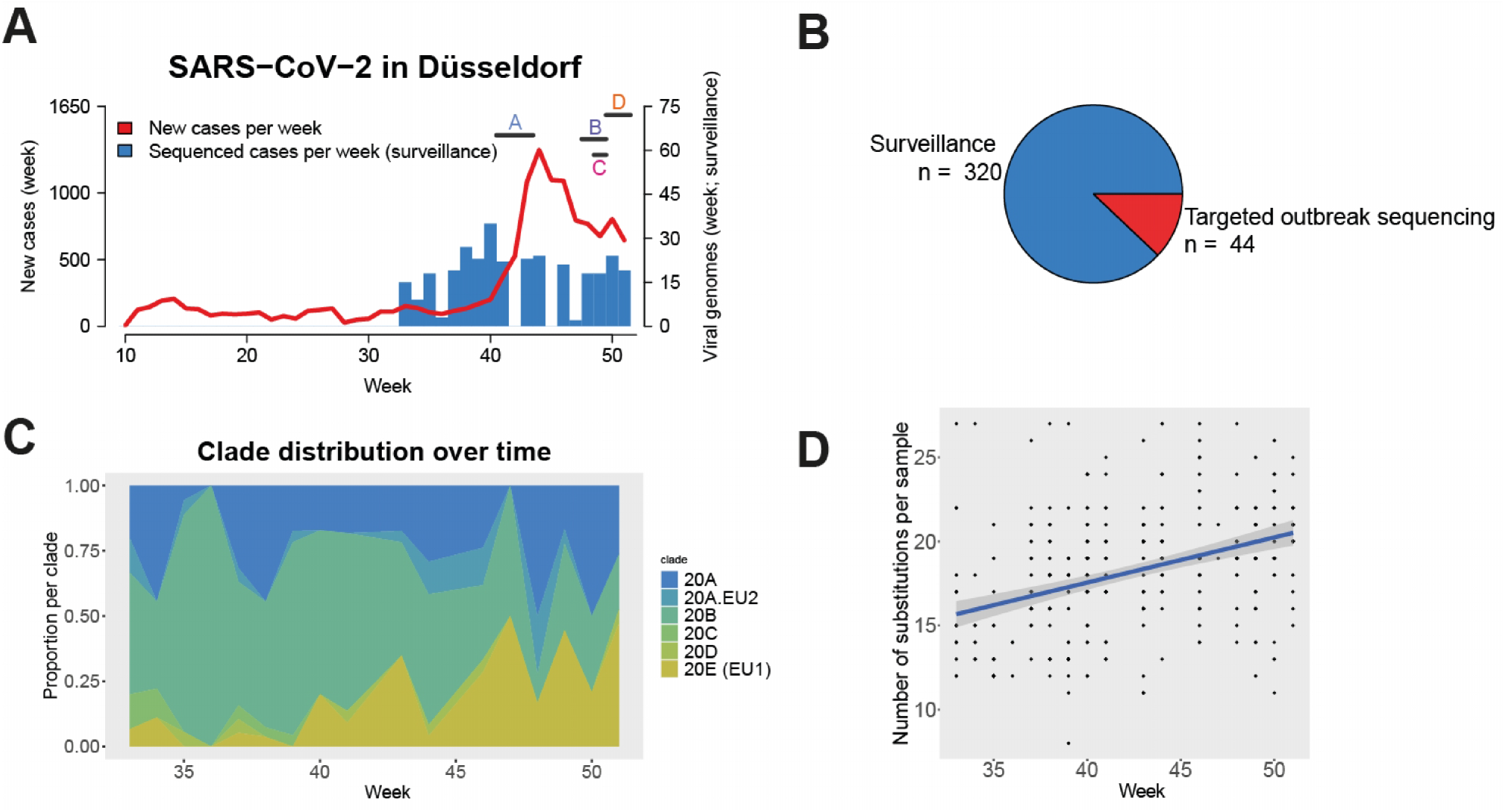
Local development of SARS-CoV-2 from September to December 2020. **A**. Newly diagnosed (red line) and sequenced (blue bars; by sample collection week) cases of SARS-CoV-2 by calendar week of 2020 in Düsseldorf. Horizontal bars indicate sample collection times for four hospital outbreaks on different wards (A – D) of Düsseldorf University Hospital. **B**. Sequenced samples by sample origin. **C**. Clade composition of surveillance samples by sample collection week, using the NextStrain [Hadfield et al. 2018] colour scheme. **D**. Substitutions per sequenced surveillance sample and sample collection line; each dot represents one viral genome, blue line: linear fit.

A comparison (see Methods; Supplementary Table 3) between the 320 surveillance samples and 385,109 non-Düsseldorf samples from GISAID [Shu and McCauley 2017] showed that 216 / 320 isolate genomes had no identical (distance = 0) or near-identical (distance = 1) sequences in GISAID, showing that many of the surveillance samples carried viral haplotypes that were not yet represented at the tip level of the global phylogenetic tree. For 104 / 320 surveillance genomes, at least one identical or near-identical GISAID sequence was found; in 29 of these cases, at least one identical GISAID sequence was found (Supplementary Table 3). A country-level analysis (see Methods) of the identified related GISAID sequences showed that sequences closely related to our surveillance samples are often found in the UK (present in the set of related sequences for 65 / 104 surveillance genomes), Switzerland (62 / 104), and Germany (41 / 104); reflecting, in general, both overall sequencing activity and geographical proximity (Supplementary Figure 1).

### Hospital Outbreak Analysis

Düsseldorf University Hospital is a maximum care facility with more than 7,000 employees, approximately 50,000 inpatients and more than 300,000 outpatients per year. After having collected surveillance data for approximately two months, we set out to use our genomic epidemiology system for the analysis of putative SARS-CoV-2 outbreaks at Düsseldorf University Hospital flagged by the hospital’s microbiology and hygiene department. Sequencing and assembly statistics on all sequenced outbreak samples are summarized in Supplementary Table 1 and Supplementary Table 2.

### Hospital Outbreak Ward A

A potential SARS-CoV-2 outbreak on ward (“A”) of Düsseldorf University Hospital was detected by the hospital’s hygiene staff in October 2020, affecting two patients and at least four healthcare workers. Using an Illumina-based sequencing strategy (see Methods), we obtained high-quality viral genomes from the two patients and four healthcare workers. A genetic analysis showed that the six sampled genomes formed a clonal cluster against the background of the collected surveillance sequencing data (Figure 3B), with inter-sample distances ranging between 0 and 1 within the set of putative outbreak samples (Figure 3C). There were no identical or near-identical (distance 0 or 1, respectively) isolates in the surveillance sequencing and GISAID datasets.

**Figure 3.**
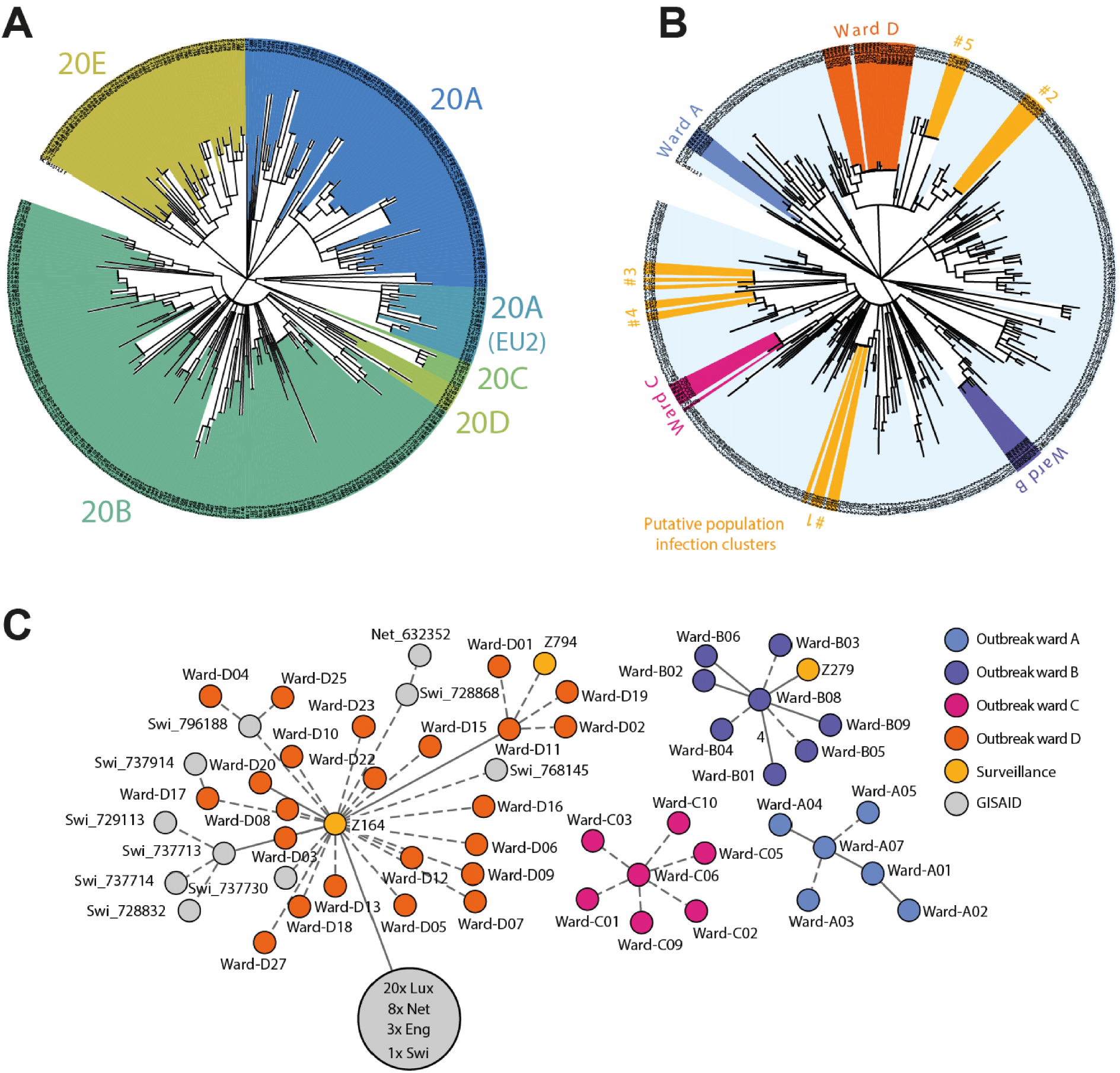
**A**. Phylogenetic tree of the 320 surveillance samples collected during this study; colours are assigned according to the NextStrain [Hadfield et al. 2018] clade system. **B**. Joint phylogenetic tree of 44 samples from four hospital outbreaks (Ward A - D) and 320 surveillance samples. For a description of the outbreaks, see main text. Putative population infection clusters are highlighted in yellow (#1 - #5). Gaps in the corresponding shaded areas correspond to related samples not identified by the greedy clustering algorithm (see Methods). Tree visualization based on iTol [Letunic and Bork 2016]. **C**. Minimum spanning tree (calculated with the Python library networkx version 2.5; visualized with Cytoscape version 3.8.2 and Inkscape version 0.92) visualization of the four hospital outbreaks, including all identical or near-identical (distance = 0 or distance = 1) from GISAID and the surveillance sequencing cohort. Samples from GISAID are labelled with their country of origin (Lux = Luxemburg; Net = Netherlands; Swi = Switzerland; Eng = England). The large grey circle represents a cluster of identical and near-identical GISAID samples. Solid lines without number indicate distance = 1 and dashed lines indicate distance = 0 between samples.

Based on these results, the presumed outbreak nature of the collected samples and the link between the infected patients and the cases of the health care workers were confirmed. It could not be conclusively established whether the index case was a patient or a health care worker.

An analysis of local conditions suggested that transmission likely took place between medical personnel during breaks. Infection control bundle measures, including re-training in the use of personnel protective equipment, were implemented to improve staff adherence to infection control rules on the ward.

### Hospital Outbreak Ward B, Outpatient Dialysis Facility

In late November, a potential outbreak on an outpatient dialysis facility ward (“B”) was detected by the hospital hygiene staff, affecting nine patients. Using Nanopore sequencing, we obtained high-quality viral genomes from eight affected individuals. These formed one cluster with seven highly related isolates (distance 0 or 1); one isolate exhibited an increased genetic distance of ≥ 4 to the other isolates (Figure 3C). A GISAID search identified no identical or near-identical samples; an analysis of the surveillance sequencing data, however, showed that there was one genetically near-identical surveillance sequencing isolate, Z279, collected approximately six weeks prior to the beginning of the putative outpatient dialysis facility outbreak. In a joint phylogenetic analysis, the putative outbreak samples clustered against the background of the surveillance sequencing samples, including the sample with an increased genetic distance (Figure 3B).

Based on these results, the eight samples were classified as one outbreak event which probably originated in Düsseldorf. No additional contact tracing to elucidate potential links between sample Z279 and the outbreak was carried out.

The settings in dialysis facilities are known to present challenging conditions for prevention of SARS-CoV-2 transmission [Meredith et al. 2020]. A post-hoc analysis of factors that may have contributed to the transmission events was carried out, and insufficient testing frequency for identification of asymptomatic infected patients, poor room ventilation, and use of medical mask-level protective equipment for patients were identified as potential factors. As a result of the investigation, ventilation conditions in the affected room were improved; patient protective equipment was upgraded to the FFP2 standard; and rapid testing with an antigen detection assay was established for every patient for every appointment, consistent with recent CDC recommendations [Centers for Disease Control and Prevention 2020].

### Hospital Outbreak Ward C

In early December, the hospital hygiene staff of Düsseldorf University Hospital identified a potential outbreak on ward “C”, affecting at least three patients and seven healthcare workers. Seven high-quality viral genomes from infected individuals were obtained by Nanopore sequencing. An analysis showed that these were genetically identical (Figure 3C) and formed a clear cluster against the background of surveillance sequences (Figure 3B). Neither identical nor near-identical isolates were identified in GISAID or in the surveillance sequencing data.

Based on these analyses, the seven samples were classified as belonging to a defined outbreak event. The outbreak potentially originated from an elderly patient requiring increased and very close attention by the hospital staff. Rapid outbreak control was achieved by an instantly initiated comprehensive screening of all patients and personnel. As a result of the investigation, measures to monitor and improve compliance with recommended routine infection prevention practices during the COVID-19 pandemic that should be applied to all patients, not just to those with suspected or confirmed SARS-CoV-2 infection, were put into place.

### Hospital Outbreak Ward D

A large-scale potential SARS-CoV-2 outbreak, affecting 16 patients and 13 healthcare workers on ward “D” of Düsseldorf University Hospital was identified by the hospital’s hygiene staff in mid-December. Using an Illumina-based sequencing strategy, we obtained 23 high-quality viral genomes from 23 infected individuals. An analysis of the generated genomes against the background of the surveillance cohort (Figure 3B) and in a minimum spanning tree (Figure 3A) showed a distinct clonal structure of the samples. Based on these results, the classification as an outbreak was established. It was not possible to establish the index case of this outbreak since several patients and staff were tested positive on the same day.

For a detailed analysis of the outbreak samples in the context of the Düsseldorf surveillance data, we split the outbreak samples into two subgroups. Subgroup-01 represented the majority viral type of the outbreak and all samples within Subgroup-01 were genetically identical; Subgroup-02 represented samples with a distance of 1 to Subgroup-01. An analysis of the surveillance data showed that the viral type of Subgroup-01 was present in the local population as early as on 1^st^ October (sample Z164; distance to majority of outbreak samples = 0) and that the viral type of Subgroup-02 was detected again in the surveillance data while the outbreak was ongoing (sample Z794, sampled on 16^th^ December). Contact tracing data collected by Düsseldorf public health authorities showed that a family member of the individual that sample Z164 was taken from was treated on another ward of the clinical department of Düsseldorf University Hospital in October 2020, establishing a possible link between the surveillance samples and the hospital outbreak on ward “D”; a SARS-CoV-2 antibody test of this family member was weakly positive for IgA. Contact tracing data also showed that a close relative of the individual that sample Z794 had been obtained from was treated on a COVID-19 ward of Düsseldorf University Hospital, to which SARS-CoV-2-positive cases from ward “D” had been transferred to. Interestingly, 5 highly related isolates, identical (distance = 0) to samples part of the ward “D” outbreak, were also identified in Switzerland (GISAID IDs 796188, 728868, 737730 768145, 737914; sampled between 9^th^ November and 28^th^ December). Near-identical samples (distance = 1) were also found in the Netherlands and England (Supplementary Table 3).

A post-hoc investigation showed that inefficient medical mask wearing as well as the fact that some patients on the ward could not wear masks for medical reasons likely contributed to the size of this outbreak. The outbreak was stopped by comprehensive screening, a temporary stop of admissions to the ward, and re-education of staff in non-pharmaceutical interventions.

### Genetically Based Identification of Population Infection Clusters

To evaluate whether the collected surveillance sequencing data could enable the genetically based identification of infection clusters and transmission chains in the population, we applied a simple greedy clustering algorithm (see Methods) to the viral sequence surveillance data, identifying groups of genetically identical samples. This analysis identified 5 putative infection clusters within the Düsseldorf samples (Figure 3B; Supplementary Table 4). Routine public health authority contact tracing data were subsequently integrated to evaluate whether these clusters reflected epidemiologically relevant associations or coincidental genetic identity.

PopClust#1 consisted of seven patient samples collected in late August; contact tracing data revealed that this cluster corresponded to a known transmission event during a school excursion in August 2020.

PopClust#2 consisted of six patient samples collected in mid-/late September; of these, three were linked via a care home, two were collected from members of the same family, and the remaining sample was linked to an otherwise unrelated primary school outbreak.

PopClust#4 and PopClust#5, consisting of four and five patient samples collected in November and December, respectively, contained samples that were linked via joint household membership, as well as samples without any obvious connections to the identified households.

Finally, PopClust#3 consisted of five samples collected in early October, of which two belonged to the same household. For two additional samples, Z132 and Z177, a reanalysis of the originally collected contact tracing data prompted by the putative genetic connection pointed to a previously unrecognized transmission chain involving Z132, Z177, and another positively tested individual X, mediated by joint membership of a martial arts gym (Z132, X) and a close personal relationship (X, Z177). Z132 and X had not identified each other as contacts, and a – not necessarily direct – link between Z132 and X was epidemiologically supported by the potential for fomite and aerosol transmission during and after training. For the remaining sample in PopClust#3, no obvious connection to the other samples was identified.

Sample ID and contact tracing information for the identified clusters are summarized in Supplementary Table 4.

### Rapid Nanopore Sequencing Experiment

To investigate the feasibility of rapid viral sequencing data generation, we measured total turnaround time for a set of 24 samples from sample receipt to bioinformatic analysis when using streamlined workflows and processes (Methods). After a total time of 28 hours (11 hours for sample and library preparation, 15 hours for sequencing, and ≤ 2 hours for bioinformatic analysis; Supplementary Figure 3), 19 of 24 genomes were resolved to high quality (<3000 N positions; Supplementary Figure 4A). Increasing the sequencing time by at least 2 hours increased the number of high-quality resolved genomes to 20; the number of resolved bases across all samples saturated after 37 hours of sequencing (Supplementary Figure 4B).

### SARS-CoV-2 Dashboard

To enable the independent interrogation of viral sequencing data by all involved stakeholders, we developed a user-friendly web application (“Dashboard”; see Methods; https://covgen.hhu.de). The dashboard is continuously updated with sequenced population and outbreak isolate genomes and enabled the targeted development of hypotheses about the genetic structure of outbreaks or transmission chains by hospital hygiene staff or local health authorities. Inter-sample genetic relatedness is visualized using a Minimum Spanning Tree (MST), and the interface supports various filtering and visualization options, e.g. enabling the targeted display of outbreaks and genetically related samples from the local viral population (Supplementary Figure 2). During the sampling period, the dashboard was a key tool to assist the interrogation of the genetic structure of the sequenced viral genomes.

## Discussion

An improved understanding of SARS-CoV-2 transmission chains is key to effective viral containment. We have shown that an integrated local SARS-CoV-2 genomic epidemiology system implemented in the state capital city of Düsseldorf has enabled the mapping of SARS-CoV-2 infection chains through hospitals and the local population. We could confirm the clonal nature of four outbreaks in a regional maximum care hospital and contribute to the design and implementation of refined infection control intervention measures, minimizing the risk of nosocomial SARS-CoV-2 transmission. We have also developed a simple algorithmic approach to identify putative infection clusters in the local population based on genetic data alone and found five such clusters in the generated surveillance data from August to December 2020. Integration with contact tracing data showed that these likely captured epidemiologically relevant viral transmission events in various settings of societal importance, such as care homes, schools, and recreational physical activities. Intriguingly, we found two potential links between viral isolates in the local population and a hospital outbreak and identified a previously unrecognized population transmission chain in a martial arts gym. Further work is required to investigate the extent to which the genetically identified clusters also encompass cryptic transmission events not captured by the existing contact tracing data, in particular for the samples for which no epidemiological link to other cluster samples could be established. Key features of our approach include the joint analysis of population and outbreak sequencing data, the integration of genetic data with contact tracing data, and the availability of a user-friendly dashboard as a central data analysis platform for all participating stakeholders.

Interestingly, we observed an increased geographical localization of SARS-CoV-2 viral variants compared to spring 2020 [Walker et al. 2020], when identical or near-identical GISAID isolates originating from geographically distant countries such as the USA or Australia were found for many samples from the Düsseldorf area. By contrast, the majority of Düsseldorf isolates characterized here were unique with respect to samples in GISAID, and when related samples were found, they typically originated from Germany or nearby European countries (Supplementary Figure 1). These findings likely reflect the increased genetic differentiation of SARS-CoV-2 genomes due to the ongoing accumulation of mutations as well as reduced international travel due to travel restrictions and lockdown measures. The resolution and accuracy of phylodynamic and phylogeographic analyses [Kühnert et al. 2016, Lemey et al. 2020, Reimering et al. 2020] will likely benefit from this increased geographical localization of individual SARS-CoV-2 viral variants. Selective sweeps of viral types with increased transmissibility [Rambaut et al. 2020], however, may counteract the observed trend of increasing geographical localization by reducing global SARS-CoV-2 genetic diversity.

When applied at scale, an integrated local genomic epidemiology system will likely contribute to a more effective management of the SARS-CoV-2 pandemic on multiple levels. First, significant uncertainties remain with respect to the relative importance of viral transmission in various settings of societal importance, such as restaurants, public transport, childcare facilities, or schools. The integration of genomic and contact tracing data for a significant proportion of infections could identify cryptic transmission events, contribute to a more quantitative understanding of transmission frequencies in different settings, and contribute to the design of improved and specifically tailored infection prevention measures. Second, as shown here, genomic epidemiology can enable the identification of large infection clusters and superspreading events in the population, which play an important role in the pandemic [Laxminarayan et al. 2020] and the containment of which is an important task for public health services. Third, genetically characterizing a large proportion of local cases may also enable locally adapted strategies for containing the spread of variants of concern [Rambaut et al. 2020]. Under the assumption that it would be desirable to sequence as high a proportion of cases as possible, a simple model calculation (see Supplementary Table 5 for an interactive Excel sheet) shows that a strategy of sequencing all positive cases would increase the total cost of the testing system by only 30 – 40%. While not insignificant, these additional costs could potentially be offset by the economic benefits of improved pandemic management; as the current costs of lockdown-like measures in many countries are significant (e.g. estimated at EUR 25-75B per week for Germany, see Florian et al. [2020]), even a small improvement could translate into overall cost-effectiveness.

Important remaining challenges include the logistics of consistently meeting the needs of local public health authorities with respect to turnaround times [Koser et al. 2012], as well as the effective interpretation and downstream integration of viral genome data. As we and many others [Greninger et al. 2015, Quick et al. 2015, Meredith et al. 2020] have demonstrated, the Nanopore technology can enable rapid turnaround times for pathogen sequencing; specifically, we achieved a sample-to-result time of 28 hours for SARS-CoV-2 in the rapid sequencing experiment. Further improvements may be possible by improved pre-analytical logistics, real-time monitoring and control of sequencing (e.g. with RAMPART; https://artic.network/rampart), and by real-time streaming of analysis results to public health authorities. The developed SARS-CoV-2 dashboard with functions for the automatic detection of candidate infection clusters could be a first step towards a more comprehensive integration of viral genome data into the routine processes of local public health authorities; further work, however, is required e.g. for the implementation of interfaces for data exchange with classical contact tracing software. In addition, the simple greedy algorithm used here to identify candidate population infection clusters could be improved by population genetics modelling [Drummond et al. 2005, Bouckaert et al. 2014] as well as by the incorporation of additional dimensions such as sampling time.

Limitations of this study include the utilized convenience sampling scheme, the relatively low proportion of sequenced positive cases over the sampling period, and the retrospective integration of genetic and contact tracing data. Addressing these in a follow-up study aiming for comprehensive case coverage, ultra-rapid turnaround times, real-time data sharing and integration with local public health authorities would be a natural extension of the work presented here and will demonstrate the full potential of integrated genomic surveillance with respect to the SARS-CoV-2 pandemic.

## Methods

### Surveillance sample collection

Surveillance sample collection was implemented in collaboration with the local diagnostic laboratory Zotz | Klimas, the largest commercial SARS-CoV-2 testing laboratory in Düsseldorf. A convenience sampling approach, arbitrarily targeting 20 – 30 samples per week, was implemented; sample selection was typically carried out on a single day and no metadata were used to determine sampling choices. Selected samples were shipped to the Institute of Virology at the Heinrich Heine University for amplification.

### Outbreak sample collection

Samples from outbreaks at Düsseldorf University Hospital were collected by local clinical staff and the employee health department and sent to the Institute of Virology at the University Hospital of Düsseldorf, which is responsible for diagnostic testing of patients and staff. All four putative SARS-CoV-2 outbreaks identified by the hospital’s hygiene staff between September to December 2020 are included in this paper. Outbreak samples were sequenced locally on the Oxford Nanopore platform or externally in collaboration with SeqIT GmbH (Kaiserslautern, Germany); see Supplementary Table 1.

### Isolation of viral genomic material

For surveillance isolates, respiratory samples from nasopharyngeal swabs were used for total nucleic acid extraction using either foodproof Magnetic Preparation Kit VI (BIOTECON Diagnostics) on KingFisher Flex (Thermo Fisher Scientific), chemagic Viral DNA/RNA 300 Kit on chemagic 360 (both PerkinElmer Inc.) or EchoLUTION Viral RNA/DNA extraction kit (BioEcho Life Sciences). For outbreak isolates. respiratory samples from nasopharyngeal swabs were used for total nucleic acid extraction using the EZ1 Virus Mini Kit v2.0 on an EZ1 Advanced XL (Qiagen, Germany) according to manufacturers’ instructions. SARS-CoV-2 was detected as described [Corman et al. 2020] with a plasmid-standard for quantification.

### Nanopore sequencing and consensus sequence generation

Viral RNA was reverse-transcribed to single-strand cDNA using random hexamers and SuperScript [Walker et al. 2019]. Viral cDNA was PCR-amplified using the Artic network SARS-CoV-2 protocol with V3 primers [Quick et al. 2017, Quick 2020, Tyson et al. 2020], employing an extended annealing/extension time of 10min. Prior to library preparation, for each sample, Artic PCR pools 1 and 2 were combined (500 ng DNA per pool). Samples that failed successful amplification were excluded from further processing. Sequencing was carried out on the Oxford Nanopore MinION device, utilizing MIN106 flow cells and the SQL-LSK109 ligation sequencing kit. Barcoding was carried out with the EXP-NBD104 and EXP-NBD114 barcoding kits.

Data analysis and generation of consensus sequences were carried out as previously described [Walker et al. 2020]. Briefly, after basecalling with Guppy v3.4.5+fb1fbfb, the Artic pipeline^1^ (see Supplementary Text 1) with default settings was applied to each sequencing run, analyzing each sample independently with Nanopolish [Loman et al. 2015] and Medaka. Generated VCF files and consensus FASTA sequences were manually curated by a) carrying out a comparison between the Nanopolish- and Medaka-based VCF files and b) visual inspection with IGV [Robinson et al. 2011], checking for i) false-positive calls; ii) polymorphic positions with more than one plausible allele; iii) false-negative calls.

### Illumina sequencing and consensus sequence generation

Libraries for sequencing on an Illumina MiSeq platform were prepared by using Illumina’s TruSeq Nano DNA Sample Preparation kit (Illumina, San Diego, USA) according to the manufacturer’s instructions.

Briefly, amplicons were purified using the Agencourt AMPure®XP system on a BioMek NX workstation (Beckman Coulter), quantified fluometric on a FluoStar Optima (BMG Labtech) using Quant-iT Picogreen dsDNA reagent (Life technologies, Darmstadt, Germany). 200 ng of input DNA from purified amplicons were used for the end-repair reaction. Adapter ligation was carried out with Illumina TruSeq Single DNA Index Sets A and B. Resulting libraries were quantified, quality checked, and normalized using an Agilent Bioanalyzer 2100. Pooled samples were loaded onto an Illumina MiSeq and sequenced with a 500 cycle MiSeq v2 reagent kit (Illumina, San Diego, USA).

Analysis of deep sequencing data was performed using Seq-IT’s *deeptype* tool, an internally developed analysis pipeline originally designed for routine anti(retro)viral drug resistance testing. In summary, the main steps of the pipeline in this case are: 1) adapter trimming from sequence reads. 2) mapping sequencing reads against the Wuhan-Hu-1 reference using the Burrows-Wheeler-Aligner (BWA) 3) generate a mpileup file and 4) computing a consensus sequence. The generated consensus sequence is then used as new reference, thereby repeating steps 2 to 4 three times. The iterations are designed for cases in which the reference sequence deviates significantly from the sample sequence (particularly in the case of deletions and insertions).

### Quality control and isolate assembly inclusion criteria

All consensus sequences with >3000 undefined (‘N’) characters were classified as low quality and excluded from all further analyses, leading to the exclusion of 21 surveillance isolate assemblies (pre-filtering: 341 surveillance assemblies; post-filtering: 320 surveillance assemblies).

### Clade assignment and variant calling

Sample clades, variants and other summary statistics were computed with the NextClade tool of NextStrain [Hadfield et al. 2018]. Pangolin (https://github.com/cov-lineages/pangolin) was used to screen for the presence of B.1.1.7.

### GISAID neighbourhood analysis

For all surveillance and outbreak isolates, closely related samples in the GISAID database [Shu and McCauley 2017] were identified based on the multiple sequence alignment (MSA) provided by GISAID (as of 25^th^ January 2021; msa_0125.tar.xz). Note that all surveillance and outbreak isolate sequences generated by us had been submitted to GISAID and were therefore part of the MSA, except for 9 of our consensus assemblies with >5% ‘N’ characters. These were manually added to the GISAID-provided alignment using mafft [Katoh and Standley 2013], using GISAID instructions for MSA extension. For a given isolate *x*, all non-Düsseldorf isolates with distance *d*(*x, y*) ≤ 1 were identified from the set of 385,109 non-Düsseldorf GISAID MSA sequences, where *d*(*x, y*) was defined as the number of differences between the MSA entries of *x* and *y*, a) ignoring leading or trailing gaps, b) accounting for IUPAC ambiguity codes, c) counting multiple subsequent non-matching gaps columns as one difference, and d) ignoring any mismatches in the MSA regions between i) the beginning of the MSA and the 20-th ACGT character of either sequence and ii) the end of the MSA and the 20 last ACGT characters of either sequence.

### SARS-CoV-2 dashboard application

The SARS-CoV-2 dashboard web application consists of a Python/Flask back-end and a browser front-end utilizing Bootstrap (https://getbootstrap.com/), JQuery (https://jquery.com/) and D3 [Bostock et al. 2011] for visualization. As a central component, the back-end maintains a distance matrix of all included isolate genomes, based on edit distances efficiently calculated with edlib [Sosic and Sikic 2017] and with the ‘N’ character configured to match all other characters. To calculate additional clonality statistics for time point *x*, the set *S*(*x*) of isolates sampled within ±7 days of *x* is determined, and clonality is defined as the proportion of genetically identical (or near-identical, with edit distance 1) unordered pairs of elements of *S*(*x*). A separate GitHub repository is used to store the included isolates’ consensus sequences and their metadata; when the repository is updated, a ‘push’ action is triggered to the server hosting the back-end with GitHub Actions, and the distance matrix is automatically updated. With both the dashboard source code and the set of included isolate genomes under version control, any analysis carried out with the dashboard is in principle fully reproducible with the GitHub commit hashes of the front- and back-end repositories and the commit hash of the sequence repository. The data flow of the dashboard is visualized in Supplementary Figure 5. All dashboard components are available under an open-source license, and instructions on how to deploy the dashboard in a CentOS7 base image are documented in the site repository.

### Phylogenetic and minimum spanning tree analyses

Phylogenetic trees were computed with RAxML [Stamatakis 2014] based on multiple sequence alignments computed with Geneious 10.2.6 (https://www.geneious.com) and visualized with iTol [Letunic and Bork 2016]. Minimum spanning trees of hospital outbreaks and related samples from the surveillance cohort and GISAID were computed with the Python library networkx version 2.5 and were visualized with Cytoscape version 3.8.2 and Inkscape version 0.92. The distance matrix for this analysis was calculated based on the same distance function used for the GISAID neighbourhood search.

### Identification of putative population infection clusters

Putative infection clusters in the surveillance sequencing data were identified by greedily clustering all isolate genomes with edit distance 0, using the dashboard distance matrix (see above), and filtering for clusters with ≥ 4 members. All candidate clusters were manually inspected.

### Integration of contact tracing data

We integrated contact tracing and case information data available at and collected by Düsseldorf Health Department (Gesundheitsamt Düsseldorf). All personally identifiable information remained at Düsseldorf Health Department.

### Fast Nanopore sequencing experiment

The following modifications were applied to the Nanopore sequencing and analysis pipeline described above to improve total turn-around time from sample receipt to generation of the consensus sequence: (i) cDNA synthesis and PCR amplification were carried out as described in the Artic network SARS-CoV-2 protocol; however, library preparation was done by two trained personnel resulting in 10.5h from sample to MinION loading (Supplementary Figure 3); (ii) live high-accuracy Guppy base calling was set up on the MinION control workstation, utilizing a GeForce RTX 2080 Ti GPU; (iii) at defined times after the start of the sequencing run (15h, 17h, 19h, 21h, 23h, 25h, 27h, 37h, 48h, 63h, 72h), the Artic bioinformatics pipeline (see above; using only Medaka; runtime 0.5 - 1.5 hours on the MinION control workstation with 40 Intel Xeon Silver 4114 cores) was applied to the generated sequencing data, and genome completeness per sample (here defined as the number of non-N bases in the generated consensus sequences) was measured. After the run had finished, final analysis was performed with the full Artic pipeline and curation of the resulting assemblies was based on both the variants called by Medaka and Nanopolish.

## Data Availability

Viral genome sequences are available on GISAID. Accessions are listed in Supplementary Table 2. All source code for the dashboard is available under the MIT license and can be downloaded from https://github.com/DiltheyLab/SARS-CoV2-Dashboard/releases/tag/v1.0

## Ethics

This study was approved by the ethics committee of the Medical Faculty of Heinrich Heine University Düsseldorf (#2020–839).

## Acknowledgements

We would like to acknowledge Jeff Barrett for discussions and for introducing the notion of clonality / heterozygosity as a useful summary statistic of surveillance sequencing data. We would like to thank the hospital hygiene staff and the employee health department medical officers for their support.

We gratefully acknowledge the Authors, the Originating and Submitting Laboratories for their sequence and metadata shared through GISAID. All submitters of data may be contacted directly via GISAID. The Acknowledgments Table for GISAID is part of the Supplement (Supplementary Table 6).

Computational support and infrastructure were provided by the “Centre for Information and Media Technology” (ZIM) at the University of Düsseldorf (Germany). Sequencing and the design of sequencing workflows were supported by the Biologisch-Medizinisches Forschungszentrum der Heinrich Heine University Düsseldorf (BMFZ).

## Deutsche COVID-19 Omics Initiative (DeCOI)

Janine Altmüller, Angel Angelov, Anna C. Aschenbrenner, Robert Bals, Alexander Bartholomäus, Anke Becker, Daniela Bezdan, Michael Bitzer, Helmut Blum, Ezio Bonifacio, Peer Bork, Nicolas Casadei, Thomas Clavel, Maria Colome-Tatche, Inti Alberto De La Rosa Velázquez, Andreas Diefenbach, Alexander Dilthey, Nicole Fischer, Konrad Förstner, Sören Franzenburg, Julia-Stefanie Frick, Gisela Gabernet, Julien Gagneur, Tina Ganzenmüller, Marie Gauder, Alexander Goesmann, Siri Göpel, Adam Grundhoff, Hajo Grundmann, Torsten Hain, André Heimbach, Michael Hummel, Thomas Iftner, Angelika Iftner, Stefan Janssen, Jörn Kalinowski, René Kallies, Birte Kehr, Andreas Keller,Oliver Keppler, Sarah Kim-Hellmuth, Christoph Klein, Michael Knop, Oliver Kohlbacher, Karl Köhrer, Jan Korbel, Peter G. Kremsner, Denise Kühnert, Ingo Kurth, Markus Landthaler, Yang Li, Kerstin Ludwig, Oliwia Makarewicz, Manja Marz, Alice McHardy, Christian Mertes, Maximilian Münchhoff, Sven Nahnsen, Markus Nöthen, Francine Ntoumi, Peter Nürnberg, Uwe Ohler, Stephan Ossowski, Jörg Overmann, Silke Peter, Klaus Pfeffer, Anna R. Poetsch, Ulrike Protzer, Alfred Pühler, Nikolaus Rajewsky, Markus Ralser, Olaf Rieß, Stephan Ripke, Ulisses Rocha, Philip Rosenstiel, Emmanuel Saliba, Leif Erik Sander, Birgit Sawitzki, Simone Scheithauer, Philipp Schiffer, Jonathan Schmid-Burgk, Wulf Schneider, Eva-Christina Schulte, Joachim Schultze, Alexander Sczyrba, Mariam L. Sharaf, Yogesh Singh, Michael Sonnabend, Oliver Stegle, Jens Stoye, Fabian Theis, Janne Vehreschild, Thirumalaisamy P. Velavan, Jörg Vogel, Max von Kleist, Andreas Walker, Jörn Walter, Dagmar Wieczorek, Sylke Winkler, John Ziebuhr

## Supplement

Supplementary Table 1

Summary of the generated sequencing data.

Supplementary Table 2

Sampling dates, GISAID accessions, clade assignments, assembly statistics, and detected variants for the sequenced samples

Supplementary Table 3

GISAID neighbourhood analysis for surveillance and outbreak samples.

Supplementary Table 4

Identified putative population infection clusters in the surveillance data and contact tracing results.

Supplementary Table 5

Interactive Excel sheet for cost calculation under the assumption that all positive cases are sequenced.

Supplementary Table 6

GISAID acknowledgements.

Supplementary Figure 1

Results of GISAID neighbourhood analysis for surveillance sequencing samples by country.

Supplementary Figure 2

Dashboard screenshot

Supplementary Figure 3

Rapid Nanopore sequencing experiment, involved steps and duration.

Supplementary Figure 4

Achieved genome completeness in the rapid Nanopore sequencing experiment; number of undefined bases by sample over time (panel A) and fraction of resolved (non-N) bases for all samples over time (panel B).

Supplementary Figure 5

Overview of data flows implemented in the developed SARS-CoV-2 dashboard web application. One GitHub repository stores the generated sequences, and separate GitHub repositories store the front- and back-end code of the web application. Updates of the sequencing data repository trigger a recomputation of the isolate distance matrix, and updates to the front-/ back-end repositories trigger a pull of the web application source code on the machine hosting the web application. Interactions between the repositories and the web application host server are implemented with GitHub Actions. For details, see Methods.

### Supplementary Text

Detailed description of the utilized variant calling and consensus generation pipeline for the analysis of Nanopore sequencing data, including commands and parameters.

## Footnotes

1 https://github.com/artic-network/artic-ncov2019.git

