## Supplementary figures and images for "Characterization of SARS-CoV-2 genetic structure and infection clusters in a large German city based on integrated genomic surveillance, outbreak analysis, and contact tracing"

### Supplementary Figure 1

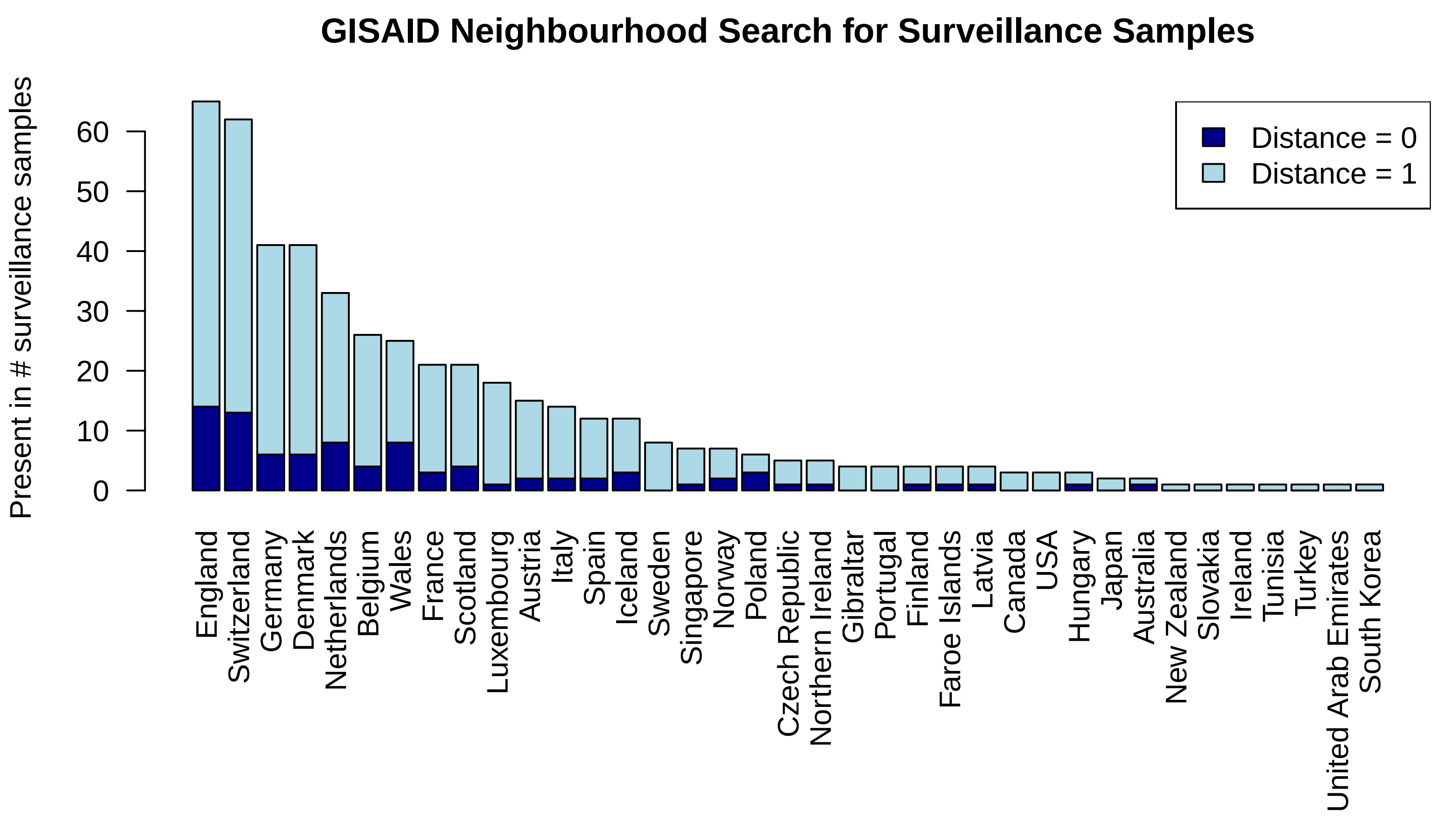

### Supplementary Figure 2

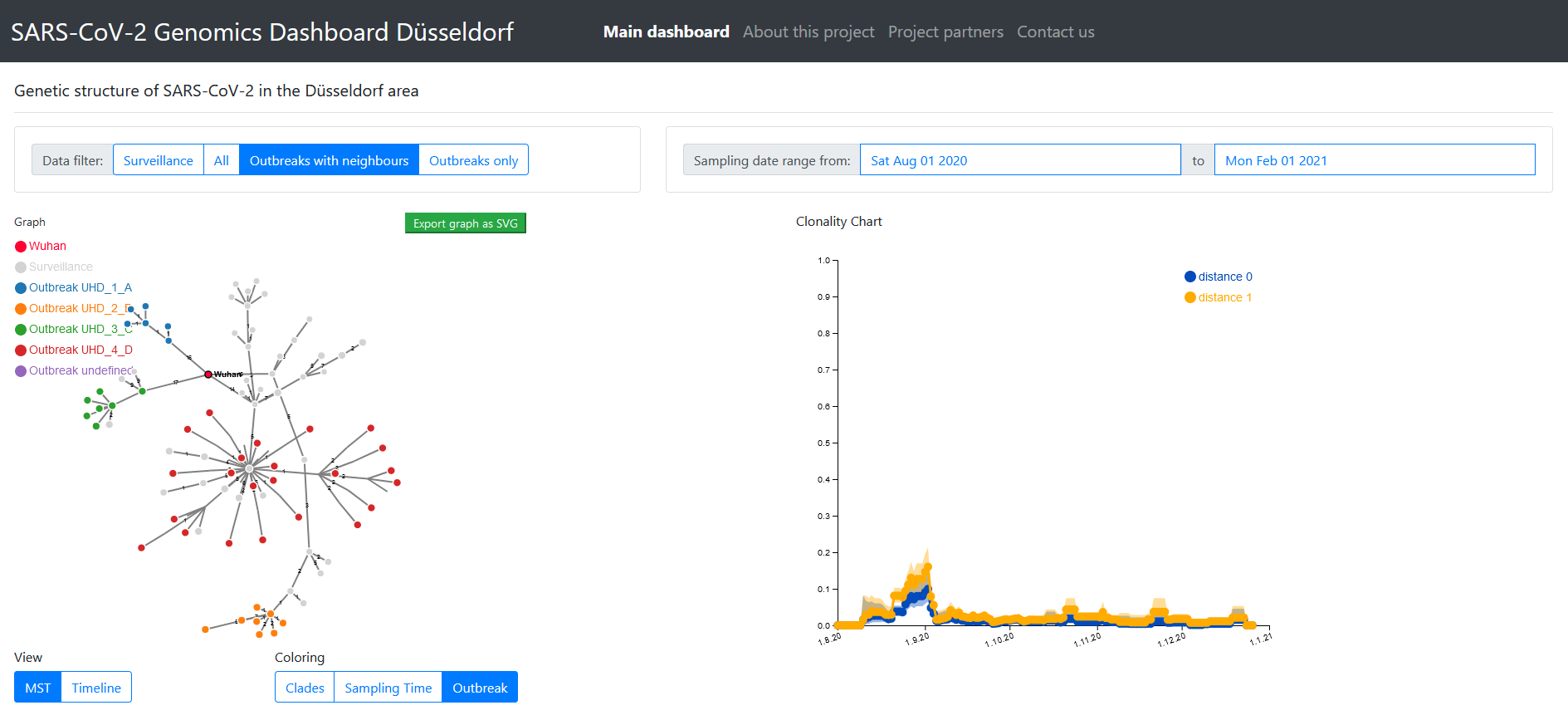

### Supplementary Figure 3

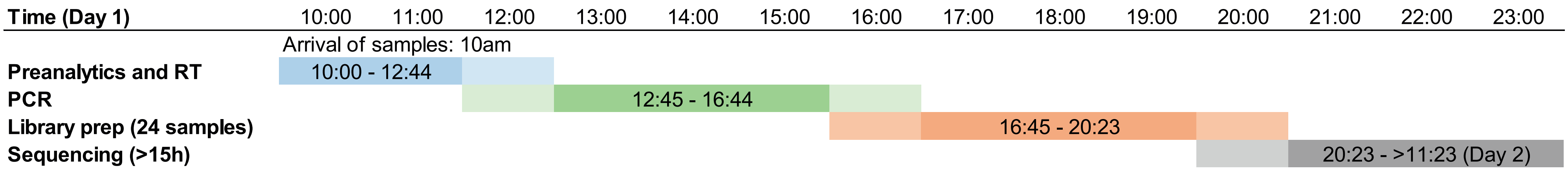

### Supplementary Figure 4

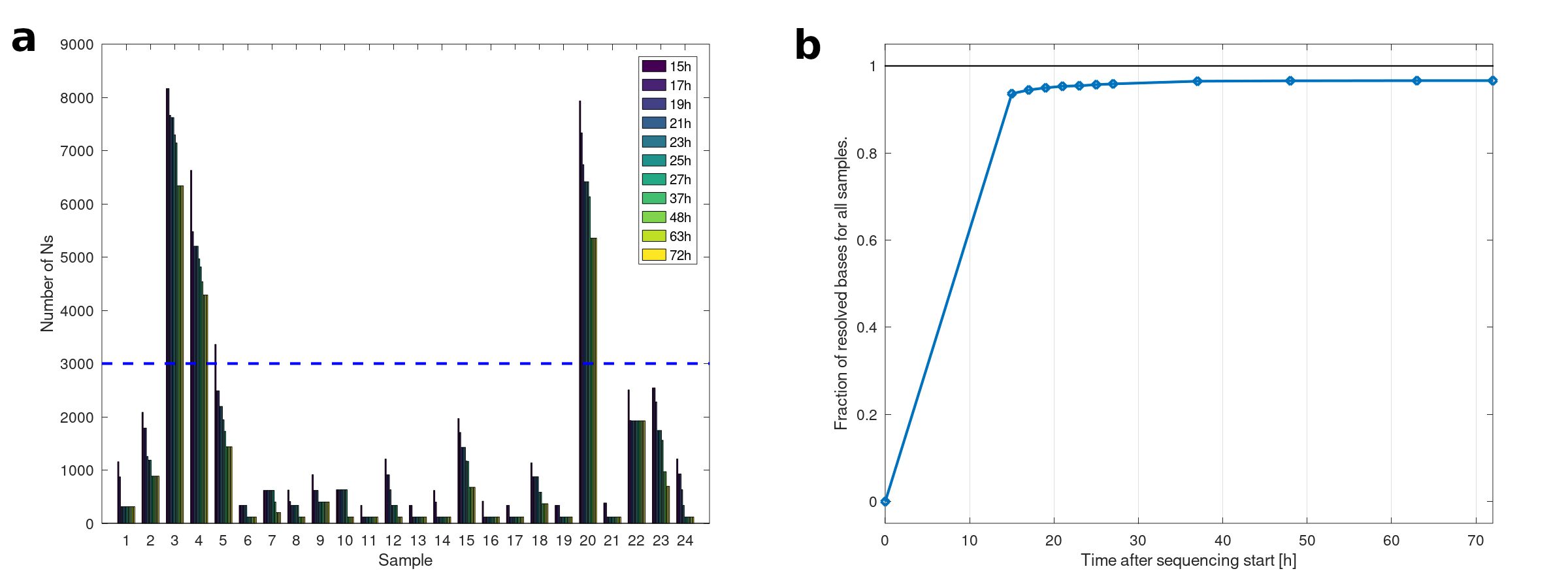

### Supplementary Figure 5

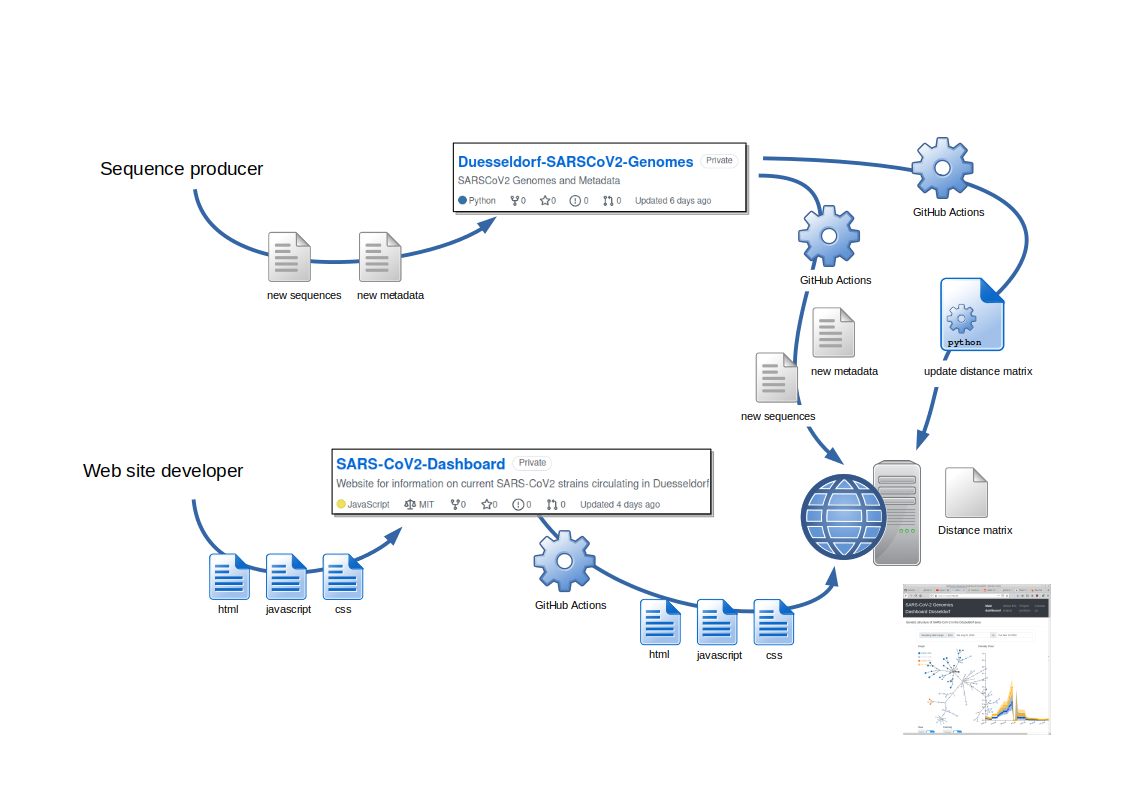
