## Supplementary Text for "Characterization of SARS-CoV-2 genetic structure and infection clusters in a large German city based on integrated genomic surveillance, outbreak analysis, and contact tracing"

### Supplementary Text 1

Consensus sequences for Nanopore sequencing data were analyzed with the Artic pipeline (<https://github.com/artic-network/artic-ncov2019.git>), comprising the following steps.

Each sample was analyzed with both Medaka and Nanopolish, followed by a round of manual curation and reconciliation of differences between the two algorithms (see main manuscript).

1. Length filtering of the generated reads to identify and remove reads that emanate from chimeric PCR fragments with artic guppyplex:

   artic guppyplex --skip-quality-check --min-length 400 --max-length 700
2. Demultiplexing with guppy_barcoder 3.4.5+fb1fbfb. The software automatically determines the utilized adapter sets und bins the reads according to their barcode:

   guppy_barcoder --recursive --require_barcodes_both_ends -i input_dir -s output_dir --arrangements_files "barcode_arrs_nb12.cfg barcode_arrs_nb24.cfg"
3. Mapping and sorting with minimap2 [Li 2018] and samtools [Li et al. 2009].

   minimap2 -a -x map-ont -t num_threads ref.fa reads.fastq | samtools view -bS -F 4 - | samtools sort -o sample.sorted.bam -
4. Coverage normalisation (target 200x) and adapter trimming with artic align_trim:

   align_trim --start –normalise 200 < sample.sorted.bam | samtools sort -T sample - -o sample.trimmed.rg.sorted.bam

   align_trim --normalise 200 --remove-incorrect-pairs < sample.sorted.bam | samtools sort -T sample - -o sample.primertrimmed.rg.sorted.bam
5. Variant calling and consensus generation with Medaka, if using the Medaka workflow:

   medaka consensus --chunk_len 800 --chunk_ovlp 400 out.hdf out.primertrimmed.medaka.vcf
   medaka variant reference.fasta out.hdf out.primertrimmed.medaka.vcf
6. Variant calling and consensus generation with Nanopolish [Loman et al. 2015], if using the Nanopolish workflow:

   nanopolish variants --min-flanking-sequence 10 -x 1000000 --progress --reads nanopolish.index -o out.primertrimmed.vcf -b out.trimmed.rg. sorted.bam -g reference.fasta -w nanopolish_header --snps --ploidy 1 -m 0.15
7. By default, genome positions with less than 20x coverage are masked with ‘N’ characters both consensus generation workflows.
